# Nationwide Spatiotemporal Dynamics and Machine Learning Prediction of Anemia Among Women in Lesotho, 2023–2024

**DOI:** 10.64898/2025.12.14.25342226

**Authors:** Tegene Atamenta Kitaw, Ribka Nigatu Haile

## Abstract

**Background:** Despite substantial efforts, anemia continues to pose a significant public health challenge, disproportionately affecting women of reproductive age. In Lesotho, insufficient data exist on its geographic distribution and key determinants using advanced techniques. This study seeks to address this gap, contributing to the development of interventions and thereby accelerating the pathway to reducing its burden.

**Methods:** This study used LDHS 2023/24 data on women of reproductive age (15–49 years), collected from 27 November 2023 to 29 February 2024 via a two-stage stratified sampling design. Spatial analysis included Global Moran’s I for autocorrelation, Getis-Ord Gi* for hotspot detection, Bayesian empirical kriging for unsampled locations, and Bernoulli-based purely spatial analysis to identify significant anemia clusters. To address class imbalance, SMOTE was applied, and feature selection was conducted using the Boruta algorithm. Multiple machine learning models were trained to predict anemia, with performance evaluated using accuracy, sensitivity, specificity, precision, F1-score, and ROC-AUC, and interpretability enhanced via SHAP method. Additionally, association-rule mining with the Apriori algorithm explored potential interactions among variables.

**Results:** The prevalence of anemia was 23.7%. Anemia exhibited significant spatial dependency (Moran’s I = 0.248, p < 0.001, Z-score = 7.774), indicating a non-random distribution. Hotspot analysis identified higher clustering in northwestern Lesotho (Leribe) and southwestern districts (Mohale’s Hoek and Quthing), while lower clustering was observed in Qacha’s Nek. The primary cluster, spanning western and central districts, represented the most pronounced high-risk area, with a relative risk of 2.34 (p < 0.001). The Gradient Boosting Machine demonstrated strong predictive performance (AUC = 87.1%), with balanced sensitivity (72.5%) and specificity (80.3%). Low education, number of children under five, poor wealth, unimproved toilet facility, occupation, urban residence, and age (35–49 years) emerged as the most influential determinants of anemia.

**Conclusions:** Anemia affects nearly one in four women in Lesotho, resulting from a complex interplay of socioeconomic, demographic, and environmental factors. Machine learning analysis, particularly the Gradient Boosting Machine, demonstrated strong predictive performance, highlighting its usefulness for predicting anemia. Effective interventions should combine geographically targeted programs in hotspot areas with broader strategies addressing women’s education, economic empowerment, sanitation, and nutrition.

## Introduction

Anemia remains a significant public health concern, affecting millions of individuals globally. Although it can occur at any age, it is particularly prevalent among women of reproductive age, with 30.7% of women aged 15–49 years affected in 2023 (1). Anemia poses a particularly severe burden in low- and middle-income countries, affecting an estimated 45.20% of pregnant women and 39.52% of non-pregnant women (2). In sub-Saharan Africa, the situation is equally alarming, with approximately 40.5% of women affected (3). Data from the World Bank indicate that the prevalence of anemia among women in Lesotho is estimated at 34% (4).

Women’s vulnerability to anemia is closely associated with both biological and social factors, including menstrual blood loss, pregnancy, nutritional deficiencies, and socioeconomic inequalities (5, 6, 7). If left unaddressed, anemia during the reproductive years can lead to serious consequences such as impaired cognitive and physical performance, reduced productivity, adverse pregnancy outcomes, and increased maternal and perinatal morbidity and mortality (8, 9, 10, 11).

A range of targeted nutrition specific interventions, including iron folic acid and vitamin A supplementation, food fortification, and programs promoting dietary diversity and food security through agricultural initiatives, have played a critical role in reducing anemia among both pregnant and non-pregnant women of reproductive age (12, 13, 14). Complementing these efforts, interventions addressing non nutritional causes, such as deworming, malaria control, and improvements in water, sanitation, and hygiene, have been strategically implemented to further mitigate the burden of anemia in this population (15, 16, 17). However, despite these comprehensive strategies, anemia remains a persistent public health challenge, indicating the need for continued and more integrated approaches.

Understanding the spatiotemporal dynamics of anemia is crucial for designing effective, targeted interventions. Traditional epidemiological methods often overlook localized variations and geographic determinants. In contrast, machine learning approaches offer powerful tools to capture complex interactions among demographic, socioeconomic, and environmental factors, providing improved predictive accuracy and actionable insights for policymakers. By combining spatial analysis with machine learning predictions, this study aims to identify high-risk clusters and generate evidence to guide context-specific interventions to reduce the burden of anemia among women of reproductive age.

## Methods

### Study Setting, Study Period, and Data Source

This study utilized data from the 2023–24 Lesotho Demographic and Health Survey (2023–24 LDHS), the fourth nationally representative DHS survey conducted in Lesotho. The survey was carried out across the entire country of Lesotho, providing comprehensive data at both national and district levels, as well as for urban and rural areas. The 2023–24 LDHS was designed to collect information to support monitoring and evaluation of the Health, Population, and Nutrition Sector Program (HPNSP) and to inform policymakers and program managers in planning and implementing health and social interventions. Data collection took place from 27 November 2023 to 29 February 2024, covering all ten administrative districts. The survey collected information on a range of sociodemographic and health indicators, including fertility, family planning, maternal and child health, childhood nutrition, and other factors relevant to monitoring the health and wellbeing of the population. The 2023–24 LDHS builds upon previous surveys to provide updated evidence on trends in health, nutrition, and social indicators in Lesotho.

### Data extraction and population

The project proposal was first submitted to the Demographic and Health Surveys (DHS) Program. After a thorough review, the DHS Program approved the proposal and issued an authorization letter granting access to the 2023–24 LDHS survey datasets. The source population included all women of reproductive age (15–49 years) in Lesotho, while the study population consisted of women within this age group living in the selected survey clusters. The authors did not have access to information that could identify individual participants during or after data collection, ensuring the confidentiality and privacy of respondents.

### Sampling methods

The 2023–24 Lesotho Demographic and Health Survey (LDHS) used the 2016 Lesotho Population and Housing Census as its sampling frame, which lists all enumeration areas (EAs) in the country. Each EA is a geographic unit, typically a city block in urban areas or a village in rural areas. The survey employed a two-stage stratified sampling design. In the first stage, 400 EAs were selected with probability proportional to size across 29 strata, including urban, peri-urban, and rural areas. A household listing was then conducted in each EA to create the sampling frame for the second stage. In the second stage, 25 households per EA were systematically selected. All women aged 15–49 who were usual members of, or slept in, the selected households the night before the survey were eligible for the Woman’s Questionnaire. All households in the subsample were eligible for the Biomarker Questionnaire. In this study, a total of 3,297 women of reproductive age were included (18).

### Variables

The primary outcome of this study was anemia status among women of reproductive age (15–49 years), classified as anemic or non-anemic based on hemoglobin levels adjusted for altitude according to WHO cut-offs (<12 g/dL for non-pregnant women, <11 g/dL for pregnant women). Blood samples were drawn from a drop of blood taken from a finger prick and collected in a microcuvette. Haemoglobin analysis was carried out on-site using a battery-operated portable HemoCue® 201+ device.

The independent variables included a range of demographic, socioeconomic, reproductive, household, and lifestyle factors. Demographic factors were age group, residence (urban/rural), ethnicity, and marital status. Socioeconomic factors included the woman’s education level, household wealth index, occupation, and husband/partner’s education. Reproductive and health-related factors comprised contraceptive use, number of births in the last five years, breastfeeding status, number of children under five, current pregnancy status, antenatal care visits, and place of last delivery. Household and lifestyle factors included type of toilet facility, smoking status, and alcohol consumption. All variables were selected based on prior evidence of their association with anemia and availability in the 2023–24 LDHS dataset. Blood samples were drawn from a drop of blood taken from a finger prick and collected in a microcuvette. Haemoglobin analysis was carried out on-site using a battery-operated portable HemoCue® 201+ device.

### Data processing and analysis

Data from the 2023–24 LDHS individual record files were extracted and processed using R software. Data cleaning involved sorting, listing, and identifying missing values, followed by weighting, recoding, and editing to ensure accuracy and consistency. Descriptive statistics, including frequencies and percentages, were calculated for all variables. Missing data were handled using appropriate imputation techniques or by excluding incomplete cases as necessary. For spatial analysis and mapping of anemia prevalence at district and regional levels, ArcGIS Pro and SatScan version 9.6 were used to visualize patterns and detect clusters. Machine learning analyses were conducted in R to predict anemia risk and identify key predictors.

### Spatial analysis of anemia

Global Moran’s I was employed to analyze spatial autocorrelation and assess the distribution of anemia among women, determining whether it was randomly distributed, clustered, or dispersed. A Moran’s I value close to “0” suggested a random distribution, a value near “–1” indicated dispersion, and a value close to “+1” signified clustering. Spatial autocorrelation was considered statistically significant if the Moran’s I p-value was below 0.05. Hot spot analysis using the Getis-Ord Gi* spatial statistic was conducted to identify statistically significant clusters of anemia. Areas with high Gi* values were classified as hot spots, indicating locations with unusually high risk of anemia, while areas with low Gi* values were classified as cold spots, indicating lower risk. These classifications help reveal spatial patterns and potential areas of concern or relative resilience.

Bayesian empirical kriging was used to estimate anemia risk in locations not directly sampled, allowing prediction of areas with higher or lower estimated risk. Purely spatial analysis using the Bernoulli model in SatScan was performed to detect statistically significant clusters of anemia, with a circular scan window representing the population at risk. Relative risk (RR) with associated p-values was used to determine the risk level of anemia within each cluster compared to areas outside the cluster.

### Machine learning analysis

The machine learning analysis of anemia among women began with data preprocessing and splitting. The dataset was randomly divided into training and testing subsets, with the training set used to develop models and the testing set reserved for evaluating performance on unseen data. Missing values were handled using imputation methods appropriate to variable type, while categorical variables were encoded using one-hot or label encoding to ensure algorithm compatibility. Numeric features were scaled using standardization or min-max normalization, depending on the requirements of the model.

To address class imbalance, which can bias machine learning models toward the majority class, several techniques were applied. Oversampling of minority cases, undersampling of majority cases, and synthetic data generation methods, such as the Synthetic Minority Over-sampling Technique (SMOTE), were used to create a balanced dataset. This process ensures that the model treats all classes equally during training, improving its ability to generalize across different population subgroups.

Feature selection was conducted using the Boruta algorithm, which iteratively compares the importance of original variables with randomized shadow features. Variables consistently demonstrating higher importance than their shadow counterparts were retained for model development, while those showing lower importance were excluded. Pairwise correlation analysis was also performed to assess multicollinearity, ensuring that the selected predictors provided largely independent information for the models.

Multiple machine learning algorithms were trained to predict anemia, including GNB (Gaussian Naïve Bayes), GBM (Gradient Boosting Machine), ANN (Artificial Neural Network), KNN (K-Nearest Neighbors), MLP (Multilayer Perceptron), SVM (Support Vector Machine), RF (Random Forest), DT (Decision Tree), XGB (Extreme Gradient Boosting). Hyperparameter tuning was performed using grid search or random search combined with cross-validation on the training set to identify the optimal parameters and reduce overfitting. Model performance was assessed using accuracy, sensitivity, specificity, precision, F1-score, and ROC-AUC, providing a comprehensive evaluation of predictive ability while accounting for class imbalance. **(*Figure 1*).**

**Figure 1:**
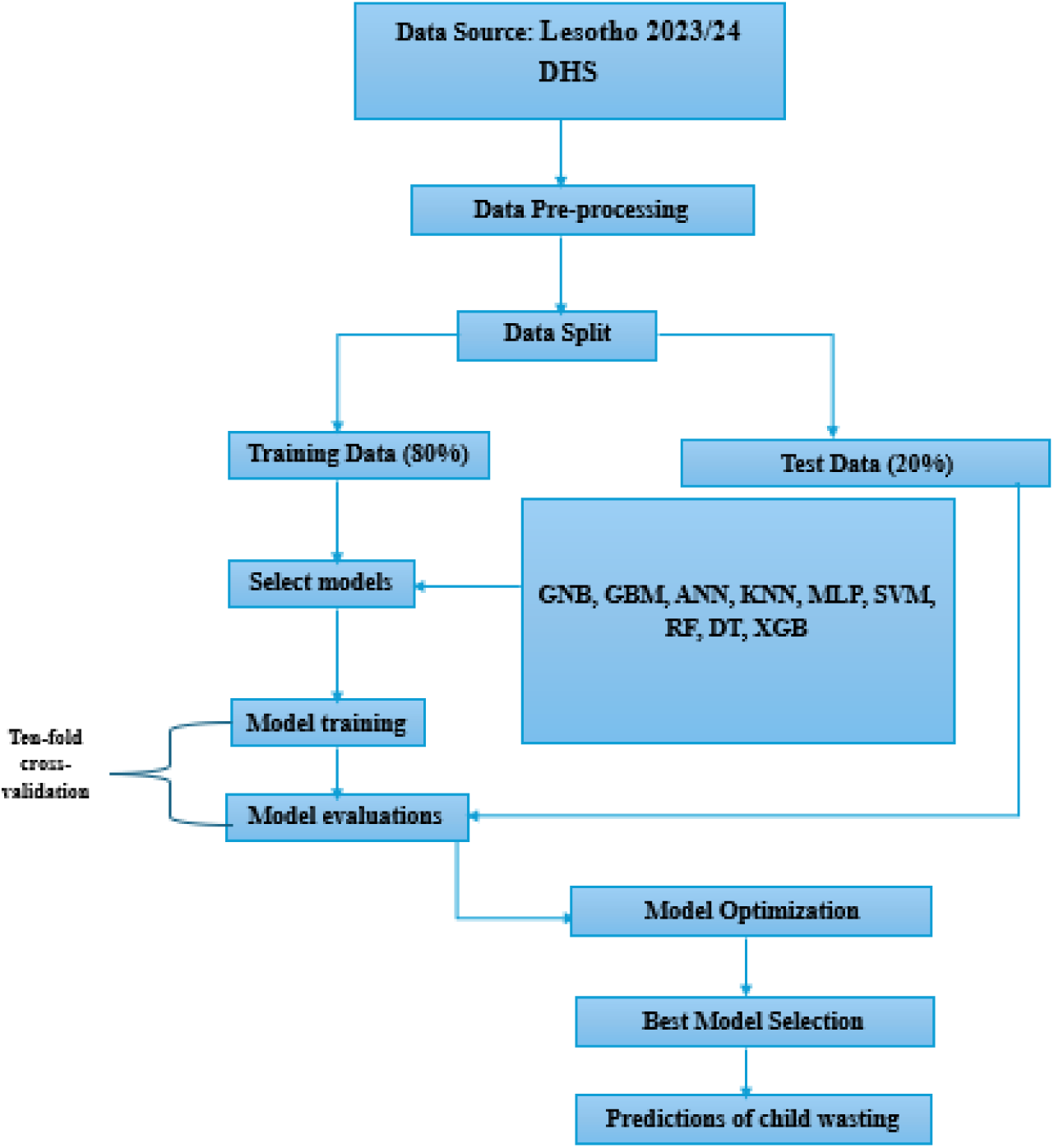
Machine learning methodology for anemia prediction in Lesotho ,2023/24.

Model interpretability was addressed using SHAP (Shapley Additive Explanations), which quantified the contribution of each feature to model predictions. SHAP values allowed the assessment of both the direction and magnitude of influence for each predictor, offering insights into the factors shaping anemia risk in a transparent and interpretable manner:

In addition, association-rule mining using the Apriori algorithm explored potential interactions among maternal, household, and child-related factors. Frequent item sets were generated, and rules were derived based on confidence and lift thresholds, systematically identifying patterns of variable combinations that could be relevant for understanding anemia risk without reporting specific outcome values.

## Results

### Descriptive statistics

The mean age of respondents was 29.28 ± 9.91 years. The study population was predominantly young women, with 39.8% aged 15–24 years, mostly of Basotho ethnicity (95.7%), and primarily living in rural areas (63.3%). Most had secondary or higher education (69.4%) and were never married (84.5%). Regarding reproductive health, 54.2% used modern contraception, and 65.3% had no births in the last five years. Additionally, 59% were not employed, 58.9% had access to improved toilet facilities, 83.2% attended four or more ANC visits, and 91.2% delivered at health facilities. ***(Table 1)*.**

**Table 1:** Descriptive statistics of women in the reproductive age group in Lesotho, 2023/24.

| Variable | Category | Frequency | Percent |
| --- | --- | --- | --- |
| Age group | 15–24 | 1,312 | 39.79 |
|  | 25–34 | 907 | 27.51 |
|  | 35–49 | 1,078 | 32.70 |
| Educational level | No education | 32 | 0.97 |
|  | Primary | 977 | 29.63 |
|  | Secondary or higher | 2,288 | 69.40 |
| Place of residence | Urban | 1,211 | 36.73 |
|  | Rural | 2,086 | 63.27 |
| Ethnicity | Basotho | 3,155 | 95.69 |
|  | Other | 142 | 4.31 |
| Marital status | Never married | 1,186 | 35.97 |
|  | Currently married/in union | 1,644 | 49.85 |
|  | Formerly married | 467 | 14.16 |
| Wealth index | Poor | 1,438 | 43.62 |
|  | Middle | 623 | 18.90 |
|  | Rich | 1,236 | 37.49 |
| Contraceptive method | No method | 1,458 | 44.22 |
|  | Traditional/Folkloric | 51 | 1.55 |
|  | Modern | 1,788 | 54.23 |
| Births in last 5 years | No births | 2,153 | 65.30 |
|  | 1 birth | 975 | 29.57 |
|  | 2+ births | 169 | 5.13 |
| Currently breastfeeding | Not breastfeeding | 2,899 | 87.93 |
|  | Breastfeeding | 398 | 12.07 |
| Children under 5 in household | No children | 1,719 | 52.14 |
|  | 1–2 children | 1,451 | 44.01 |
|  | 3+ children | 127 | 3.85 |
| **Type of toilet facility** | Improved | 1,942 | 58.90 |
|  | Unimproved | 1,355 | 41.10 |
| **Smoking status** | Non-smoker | 3,224 | 97.79 |
|  | Smoker | 73 | 2.21 |
| **Alcohol consumption per day** | No drink | 2,402 | 72.85 |
|  | 1–2 drinks | 263 | 7.98 |
|  | 3+ drinks | 632 | 19.17 |
| **Currently pregnant** | Not pregnant | 3,207 | 97.27 |
|  | Pregnant | 90 | 2.73 |
| **Husband/partner's education** | No education | 189 | 11.77 |
|  | Primary | 681 | 42.40 |
|  | Secondary/Higher | 736 | 45.83 |
| **Respondent's occupation** | Not Employed | 1,946 | 59.02 |
|  | Employed | 1,351 | 40.98 |
| **Number of ANC visits** | No visit | 34 | 4.20 |
|  | 1–3 visits | 102 | 12.61 |
|  | 4+ visits | 673 | 83.19 |
| **Place of delivery** | Home | 71 | 8.78 |
|  | Health facility | 738 | 91.22 |

## Spatial dynamics of anemia among women

### Spatial autocorrelation

The spatial distribution of anemia among women in Lesotho is clustered, with a Global Moran’s I value of 0.248, a p-value < 0.001, and a Z-score of 7.774. This indicates that anemia exhibits spatial dependency. Furthermore, the likelihood of this clustered pattern occurring by random chance is less than 1%. **(*Figure 2*).**

**Figure 2:**
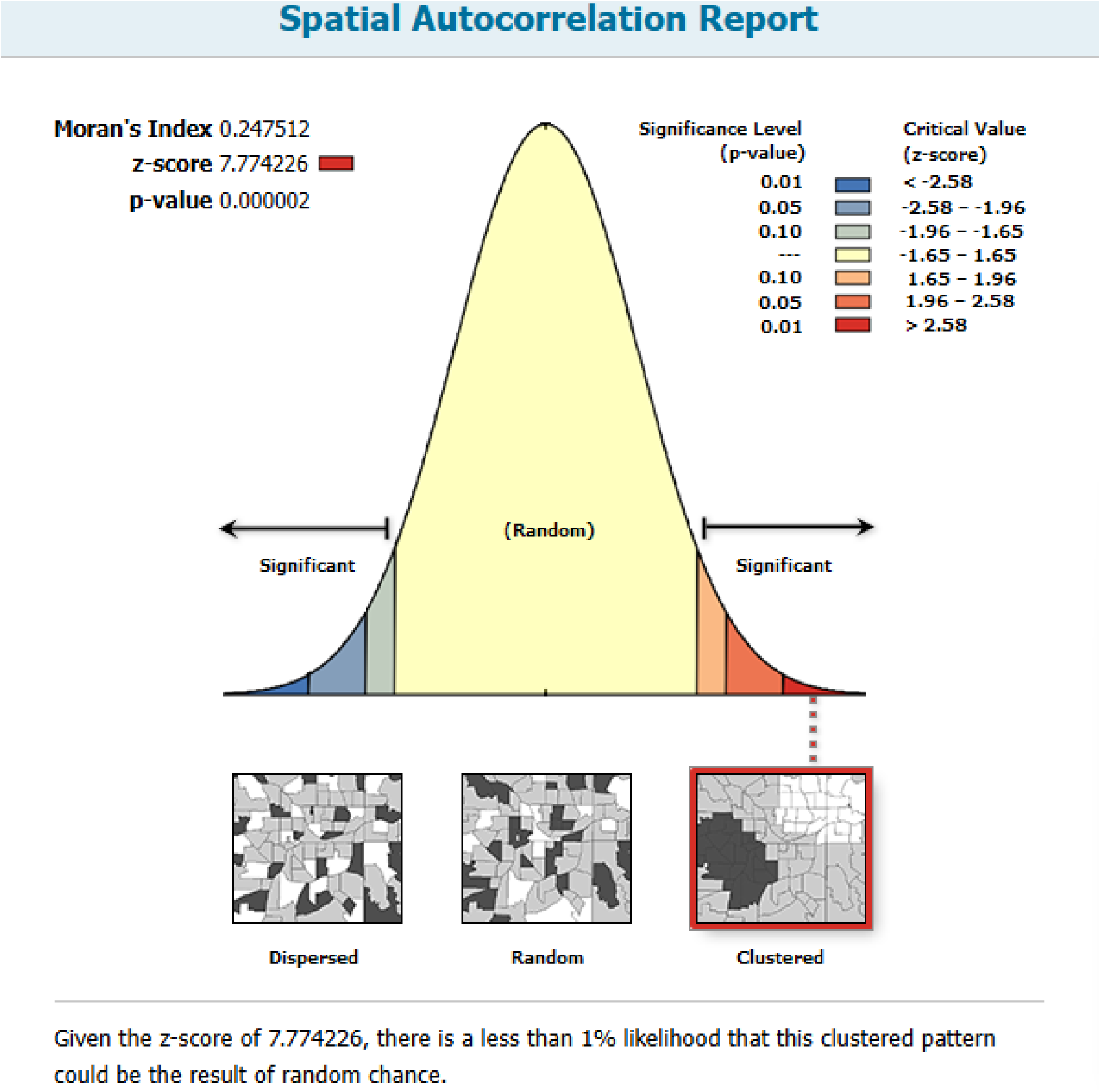
Spatial autocorrelation of anemia among women in Lesotho, 2023/24.

### Hotspot analysis

Hotspot and cold spot analyses were conducted to identify geographic areas with high and low prevalence of anemia. The results revealed significant hotspot of anemia in the northwestern region of Lesotho, in Leribe, as well as in the southwestern region of Lesotho, in Mohale’s Hoek and Quthing. In addition, cold spot of anemia was found in the eastern part of Lesotho, in Qachas Nek. **(*Figure 3*).**

**Figure 3:**
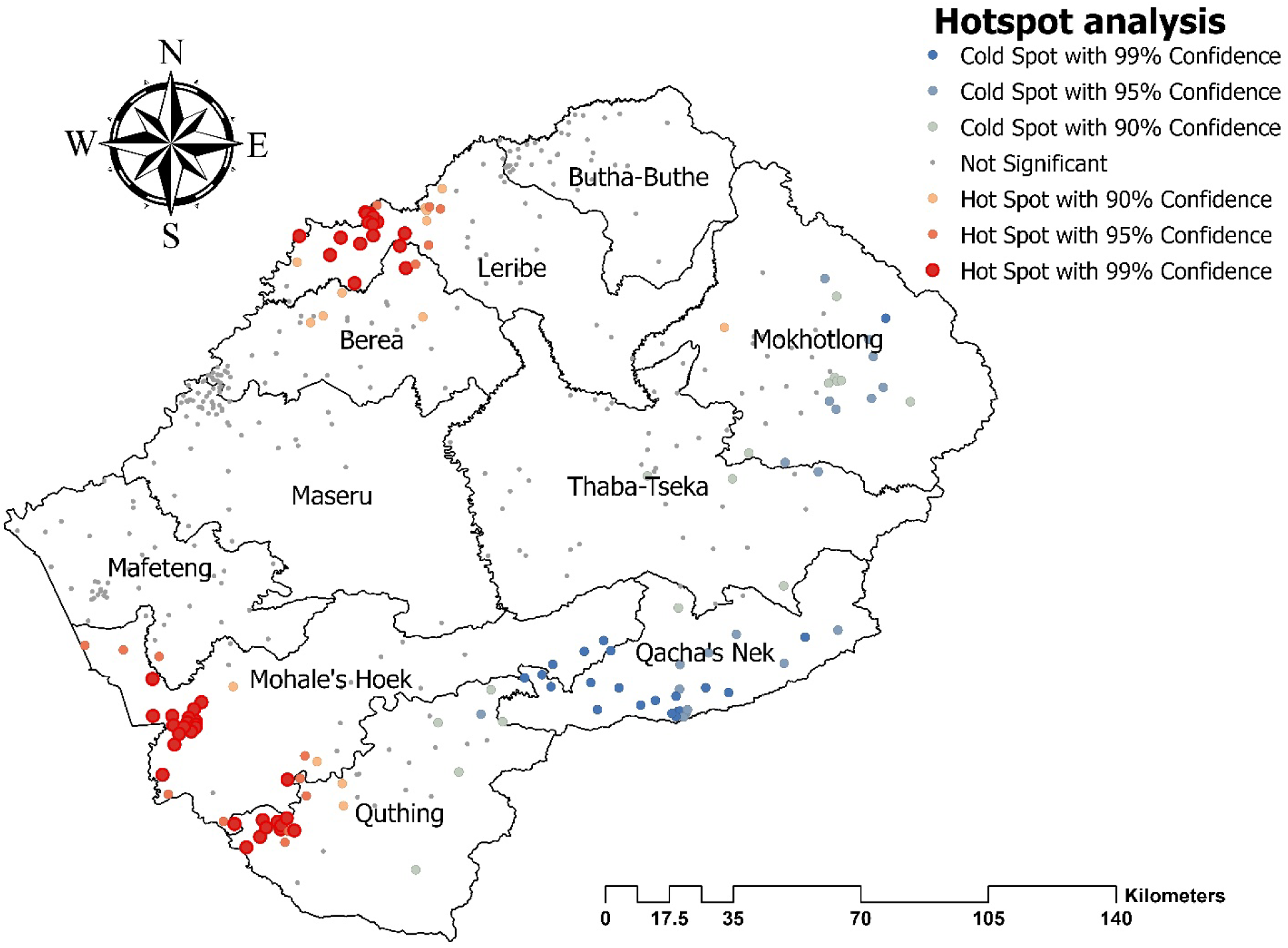
Hotspot analysis of anemia distribution among women in Lesotho, 2023/24.

### Spatial interpolation

The Empirical Bayesian Kriging analysis predicted the spatial distribution of anemia among women across Lesotho. The results indicate that the highest predicted prevalence of anemia is concentrated in the northern and southern regions. Specifically, the northern district of Leribe and the southern districts of Mohale’s Hoek and Quthing show the most elevated predicted values, highlighted in red and orange shades on the map. Moderate prevalence is observed in central regions such as Maseru, Thaba-Tseka, and Berea, represented by yellow and light green colors. In contrast, the eastern parts of the country, particularly Qacha’s Nek and Mokhotlong, show the lowest predicted prevalence of anemia, indicated by blue colors. **(*Figure 4*).**

**Figure 4:**
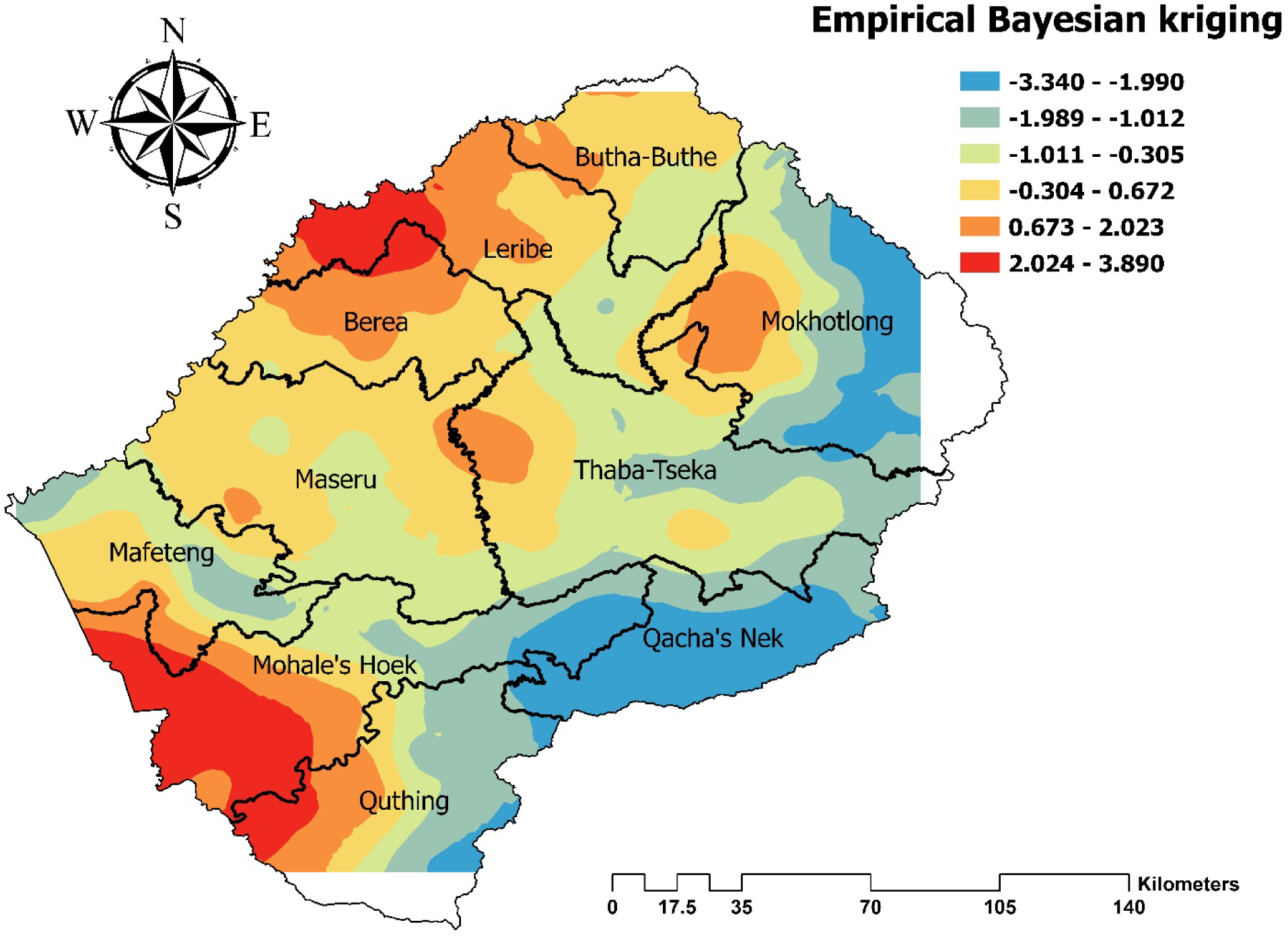
Empirical Bayesian Kriging interpolation to predict the distributions of anemia among women in Lesotho, 2023/24.

### Satscan analysis

The SaTScan analysis identified significant spatial clustering of anemia among women across Ethiopia. The results revealed that the western and central districts, including Berea, Maseru, Mafeteng, and Mohale’s Hoek, formed the most pronounced high-risk cluster, with a relative risk (RR) of 2.34 (p < 0.001), coordinates/radius: (29.543805° S, 27.257749° E) / 100.41 km, indicating that women in these areas are more than twice as likely to experience anemia compared to the national average. A secondary cluster of moderate risk was detected in the eastern district of Mokhotlong, with a relative risk of 1.37 (p < 0.001), coordinates/radius: (29.236126° S, 28.890966° E) / 4.84 km, suggesting an elevated but comparatively lower likelihood of anemia. Several districts, such as Leribe, Butha-Buthe, Thaba-Tseka, Qacha’s Nek, and Quthing, were not part of any significant clusters, indicating no statistically notable increase in anemia risk. (**Figure 5).**

**Figure 5:**
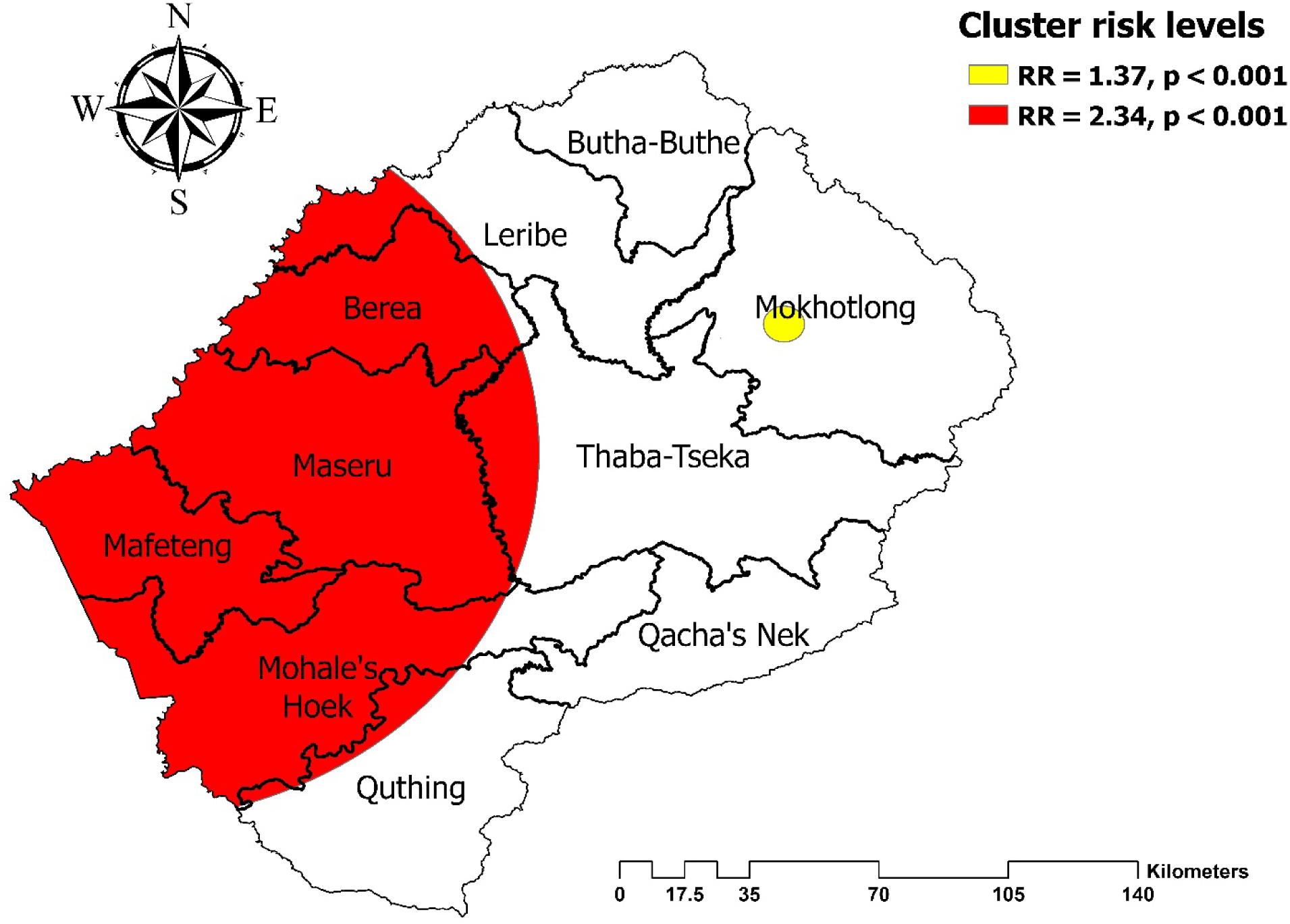
Spatial Satscan analysis of anemia among women in Lesotho, 2023/24.

## Machine learning analysis of anemia among women

### Data balancing

In the dataset, 2,515 individuals (76.3%) were non-anemic, while 782 individuals (23.7%) were anemic, indicating an imbalanced class distribution. Such imbalance can cause machine learning models to be biased toward the majority class (non-anemic), reducing their ability to correctly predict the minority class (anemic). To address this, data balancing techniques—such as oversampling the anemic cases, under sampling the non-anemic cases, or using synthetic data generation methods like SMOTE—were applied. After balancing, the dataset contained roughly equal proportions of anemic and non-anemic individuals, with each class representing approximately 50% of the total samples. **(*Figure 6*).** This adjustment helps to ensure that the model treats both classes equally, improving its ability to accurately identify anemic cases.

**Figure 6:**
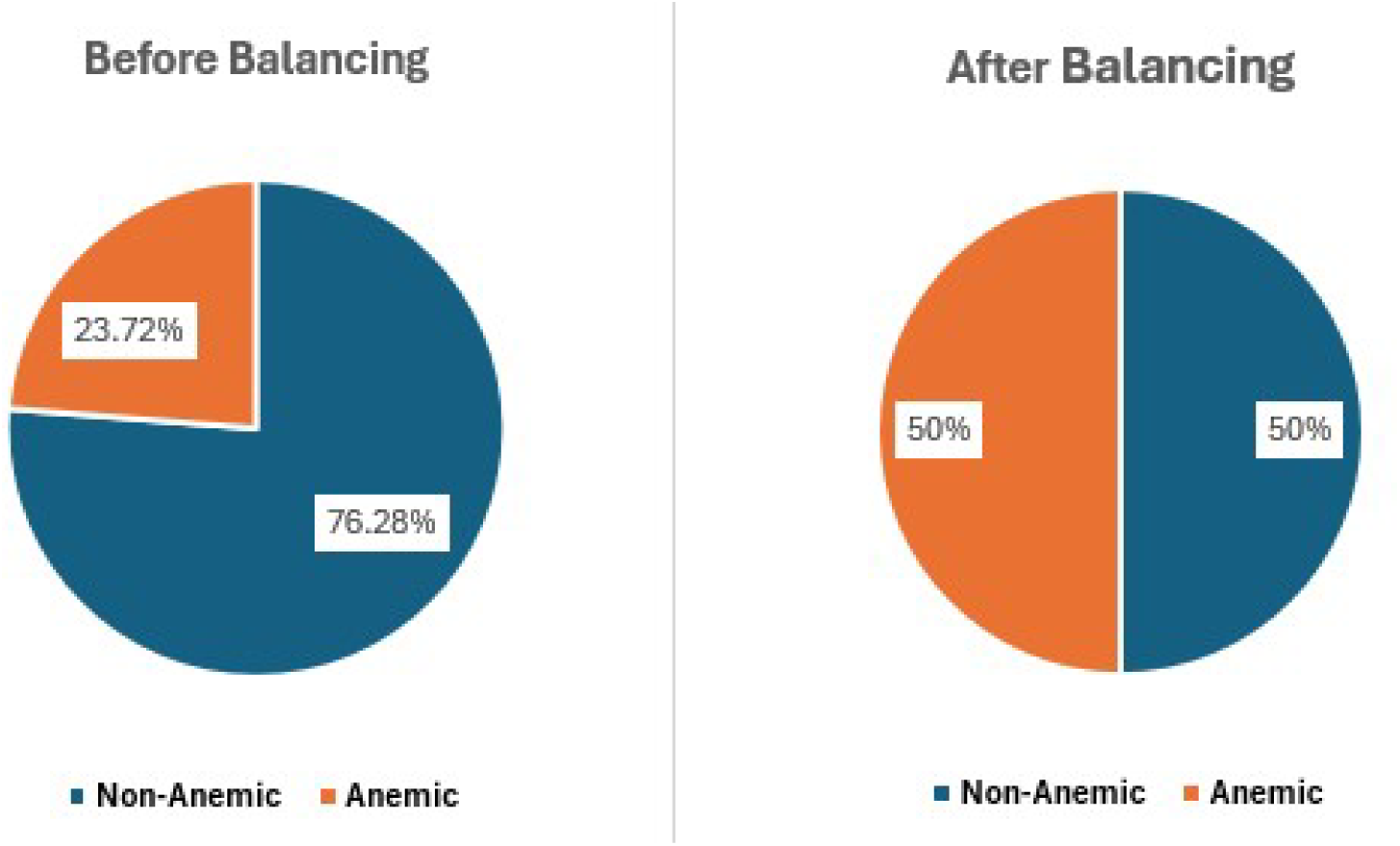
Distribution of anemic and non-anemic women before and after data balancing.

### Feature selection

The Boruta algorithm was applied to identify the most relevant features influencing anemia status. By iteratively comparing the importance of original variables with that of randomized shadow features, Boruta classifies variables as either confirmed or rejected. Confirmed variables, which consistently demonstrated higher importance than their shadow counterparts, were considered key contributors to variations in anemia and retained for further modeling. Rejected variables, which failed to show significant importance, were excluded from subsequent analysis. In the Boruta output, confirmed variables are highlighted in green and rejected variables in red (19). Using this approach, eight features were identified as significant predictors of anemia: educational level, place of residence, wealth index, contraceptive method, births in the last five years, number of children under five in the household, type of toilet facility, and respondent’s occupation. **(*Figure 7*).**

**Figure 7:**
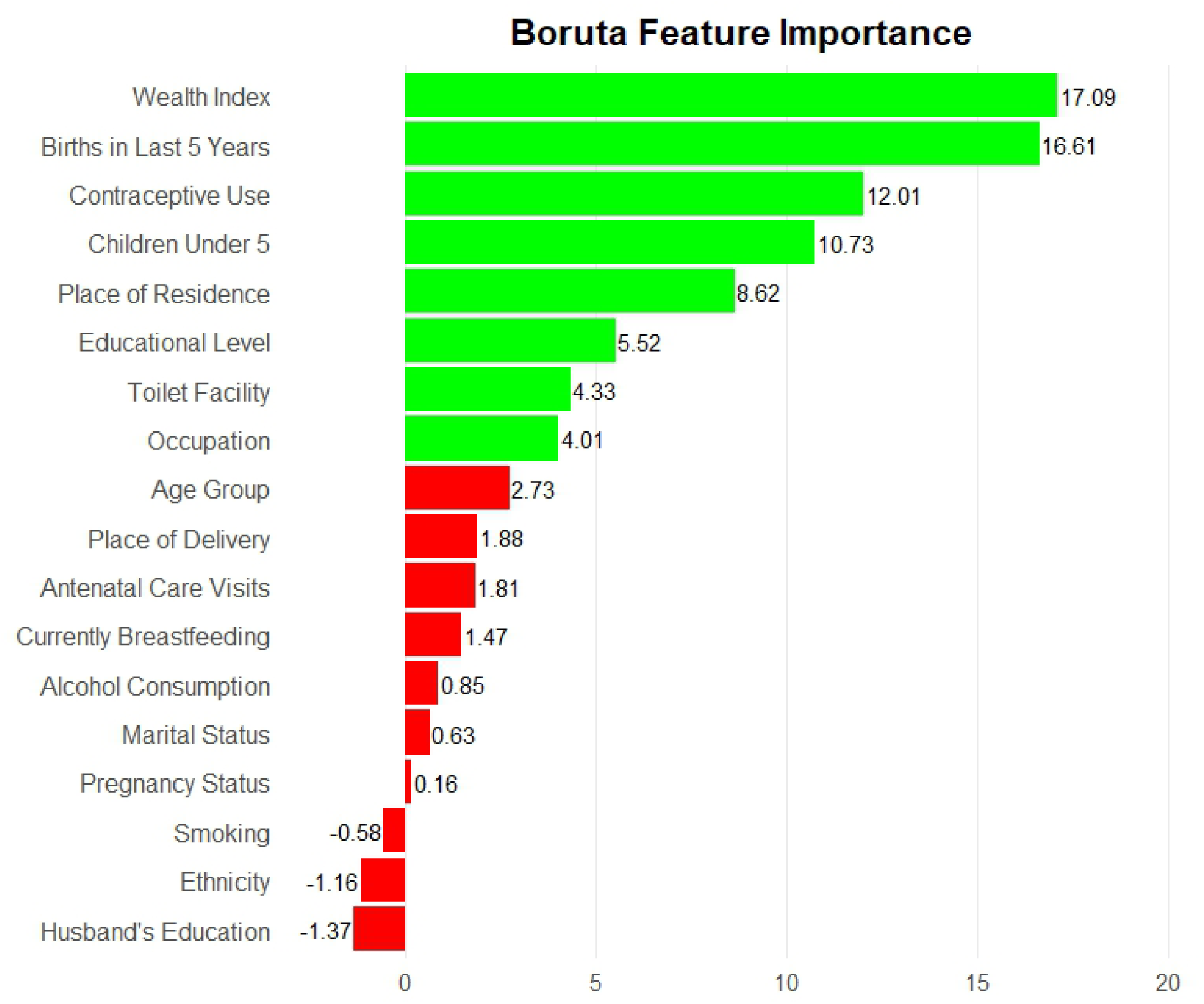
Boruta algorithm identifying the most relevant features influencing anemia status in Lesotho, 2023/24.

The heat map of the correlation matrix displays the pairwise relationships among all features in the dataset. Each cell represents the correlation coefficient between two features, with color intensity indicating the strength and direction of the correlation. In this case, all correlations are below 0.8, indicating that no pairs of features are highly correlated. This suggests minimal multicollinearity, meaning the features provide largely independent information, which is favorable for predictive modeling and ensures that each variable can contribute uniquely to the analysis. (***Figure 8***).

**Figure 8:**
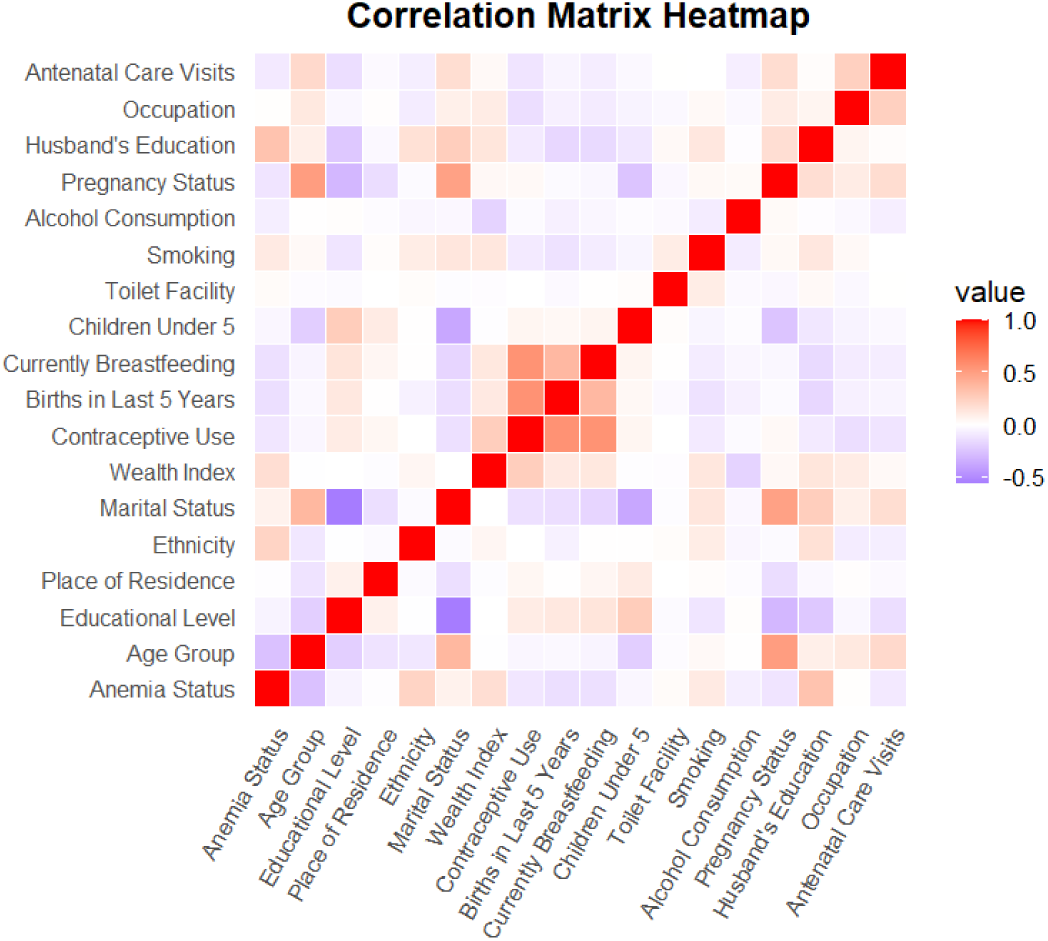
Heat map showing the pairwise correlations among all features.

The performance of multiple machine learning models in predicting anemia is summarized in the table. Among the models evaluated, Gradient Boosting Machine (GBM) achieved the highest overall performance, with a sensitivity of 72.5%, specificity of 80.3%, precision of 68.0%, accuracy of 83.5%, F1-score of 70.2%, and an ROC-AUC of 87.1%, indicating strong discriminative ability.

Extreme Gradient Boosting (XGB) and Random Forest (RF) also performed well, with accuracy above 70% and ROC-AUC above 80%, demonstrating their robustness in capturing complex relationships in the data. Models such as Gaussian Naive Bayes (GNB) and Artificial Neural Network (ANN) showed lower predictive performance, with accuracies around 53–61% and ROC-AUC below 63%, suggesting limited effectiveness in distinguishing anemic from non-anemic individuals. Overall, tree-based ensemble models, particularly GBM and XGB, outperformed other algorithms, highlighting their suitability for predicting anemia based on the selected features. (***Table* 2**). In addition, based on the ROC curve analysis, the top three machine learning algorithms for classifying anemia were GBM (AUC = 0.87), XGB (AUC = 0.84), and Random Forest (AUC = 0.83), indicating their superior discriminative ability compared to the other models.(***Figure 9***).

**Figure 9:**
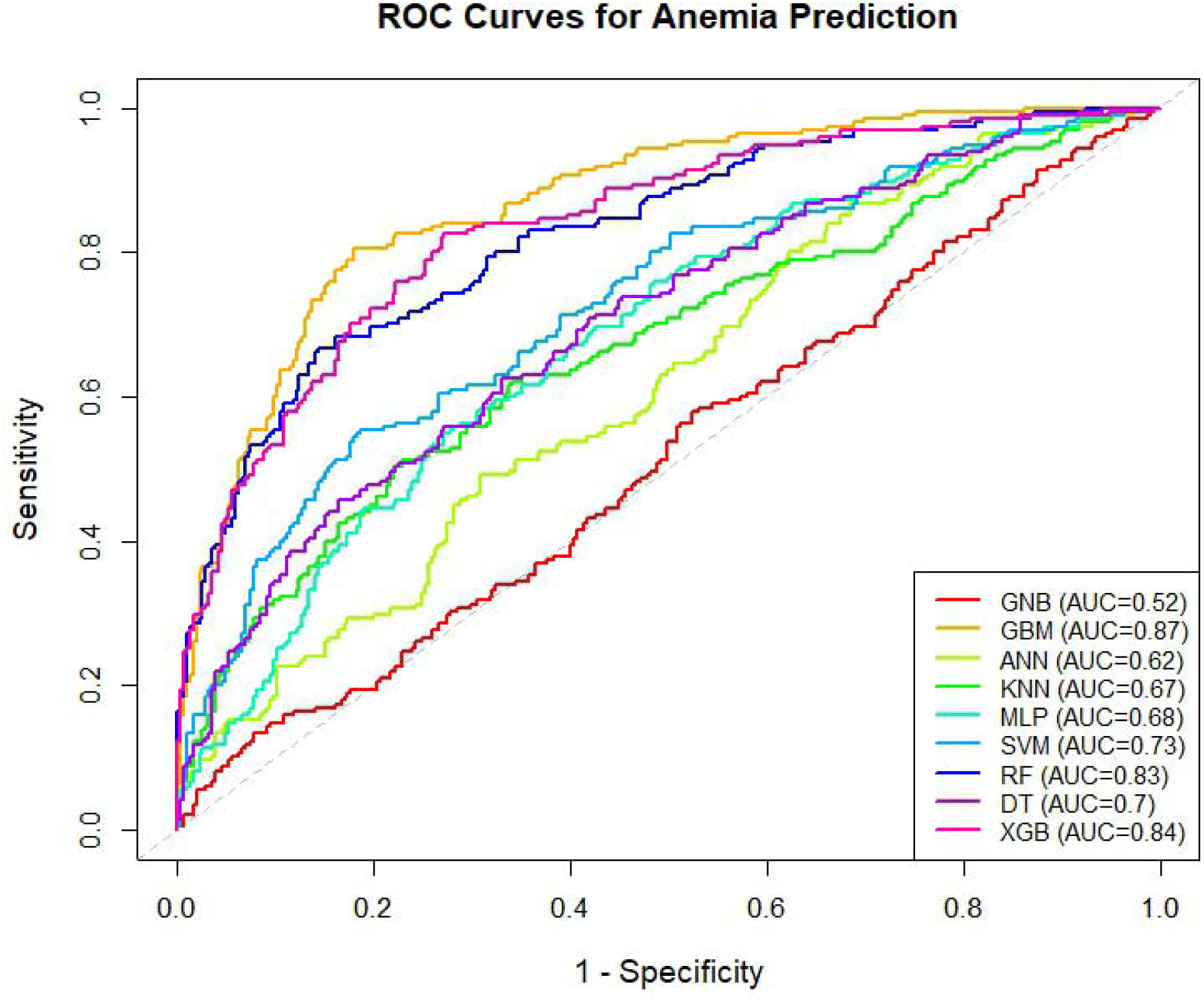
ROC curve analysis of the top-performing machine learning models.

**Table 2:** Performance Metrics of Machine Learning Models for Predicting Anemia.

| Model | Sensitivity (%) | Specificity (%) | Precision (%) | Accuracy (%) | F1_score (%) | ROC (%) |
| --- | --- | --- | --- | --- | --- | --- |
| GNB | 50.2 | 56.0 | 27.5 | 53.0 | 35.8 | 52.3 |
| GBM | 72.5 | 80.3 | 68.0 | 83.5 | 70.2 | 87.1 |
| ANN | 60.1 | 62.3 | 45.0 | 61.0 | 51.7 | 62.4 |
| KNN | 55.3 | 67.5 | 50.2 | 61.5 | 52.7 | 67.0 |
| MLP | 65.0 | 58.0 | 48.5 | 61.5 | 55.8 | 68.3 |
| SVM | 58.2 | 70.0 | 52.4 | 64.0 | 55.1 | 73.0 |
| RF | 68.0 | 75.5 | 61.5 | 71.8 | 64.5 | 83.4 |
| DT | 60.5 | 65.0 | 50.0 | 62.5 | 54.9 | 70.0 |
| XGB | 70.0 | 78.0 | 65.2 | 73.5 | 67.5 | 84.4 |

## Model interpretability

### SHAP plot interpretation

Education levels (yellow points) are associated with negative SHAP values, suggesting a protective effect, while lower education (purple) shifts predictions toward higher anemia risk. Wealth index also shows a strong protective role, with wealthier women less likely to experience anemia. Age contributes moderately, where older women (yellow) tend to have slightly higher anemia risk compared to younger women. Births in the last 5 years increase the likelihood of anemia, as higher values are associated with positive SHAP contributions. Finally, place of residence has the smallest effect, with urban residence showing a modest protective influence relative to rural residence. Overall, the model highlights that socioeconomic status (education and wealth) and reproductive history are the dominant factors shaping anemia risk, with age and residence playing smaller roles. (***Figure 10***).

**Figure 10:**
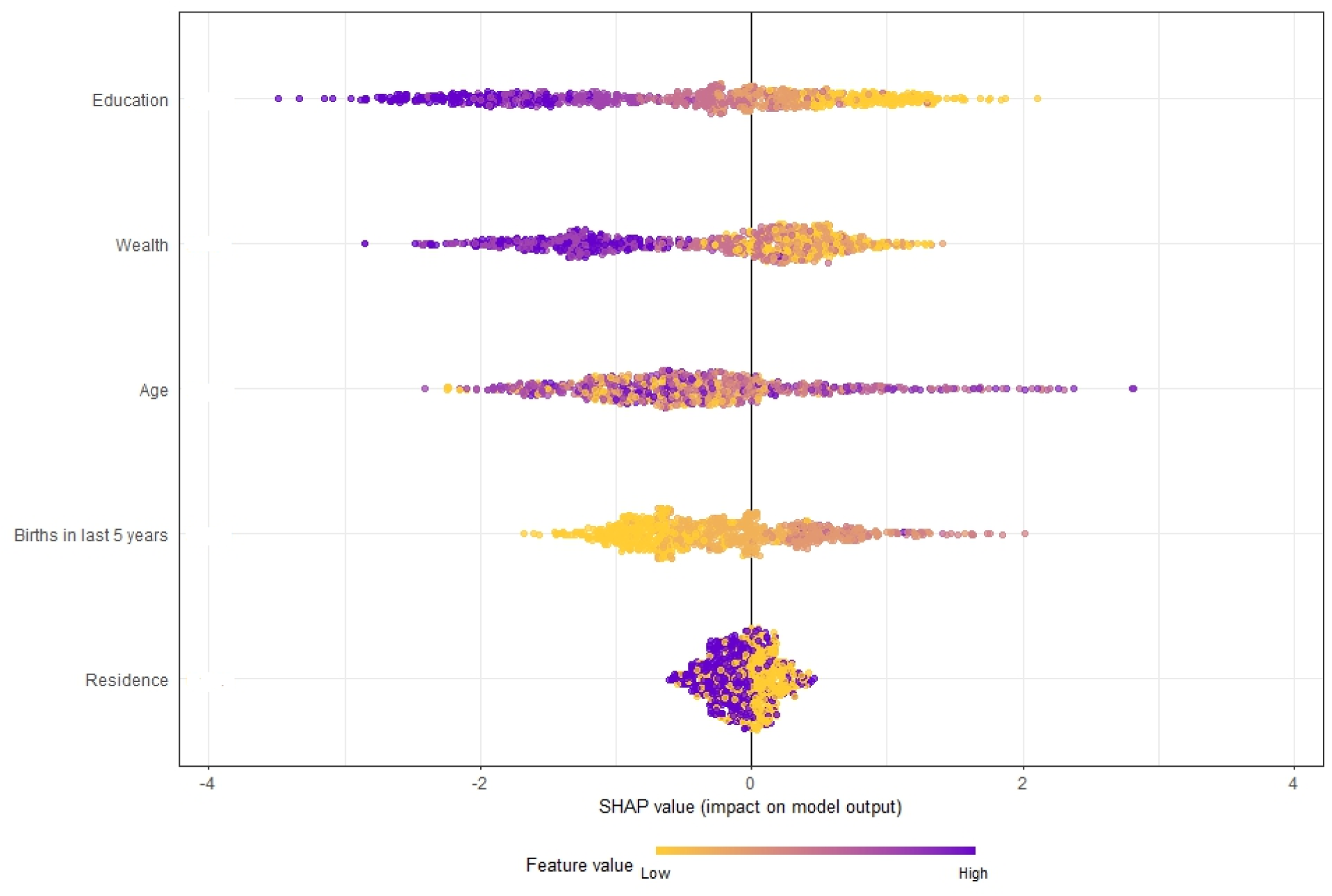
SHAP plot showing feature contributions to anemia risk.

### Association Rule Mining

By employing the Apriori algorithm, our study systematically explored patterns and combinations of maternal, household, and child-related factors associated with anemia among women. The analysis revealed several association rules that provide insight into how different factors interact to influence anemia risk. Key variables that repeatedly appeared in the top rules include maternal education, number of children under 5 in the household, household wealth, toilet type, and maternal occupation, highlighting the multifactorial nature of anemia risk among women.

Women in households with 1–2 children under 5 had a probability of anemia of approximately 79% (confidence = 0.79, lift = 1.18), indicating that the presence of young children is a common factor across the top rules. Women with lower maternal education levels who also lived in households with 1–2 children under 5 had a probability of anemia of 76% (confidence = 0.76, lift = 1.18), emphasizing the role of educational attainment in anemia risk. In households classified as poor, women with 1–2 children under 5 had a probability of anemia of 81% (confidence = 0.81, lift = 1.18), showing that lower socioeconomic status contributes significantly to increased risk. Household sanitation also appeared as an important factor: women living in households with 1–2 children under 5 and an unimproved toilet type had a probability of anemia of 78% (confidence = 0.78, lift = 1.18). Maternal occupation showed a probability of 75% (confidence = 0.75, lift = 1.18) in the same group. Notably, the combination of low maternal education, poor household wealth, and 1–2 children under 5 produced the highest probability of anemia at 82% (confidence = 0.82, lift = 1.18).

### Evaluation of Feature Relevance

We used the Gradient Boosting Machine (GBM) algorithm to analyze the importance of features in predicting anemia among women. Choosing GBM over SVM, despite their comparable predictive performance in this study, was due to GBM’s interpretability advantage. Unlike SVM, GBM inherently provides feature importance scores, allowing us to directly identify which variables most strongly influence anemia risk.

Accordingly, the top seven most important variables for predicting anemia in this study were maternal education, number of children under five in the household, household wealth, toilet type, maternal occupation, residence type, and age group. These variables collectively highlight the multifactorial nature of anemia risk among women. **(*Figure 11*).**

**Figure 11:**
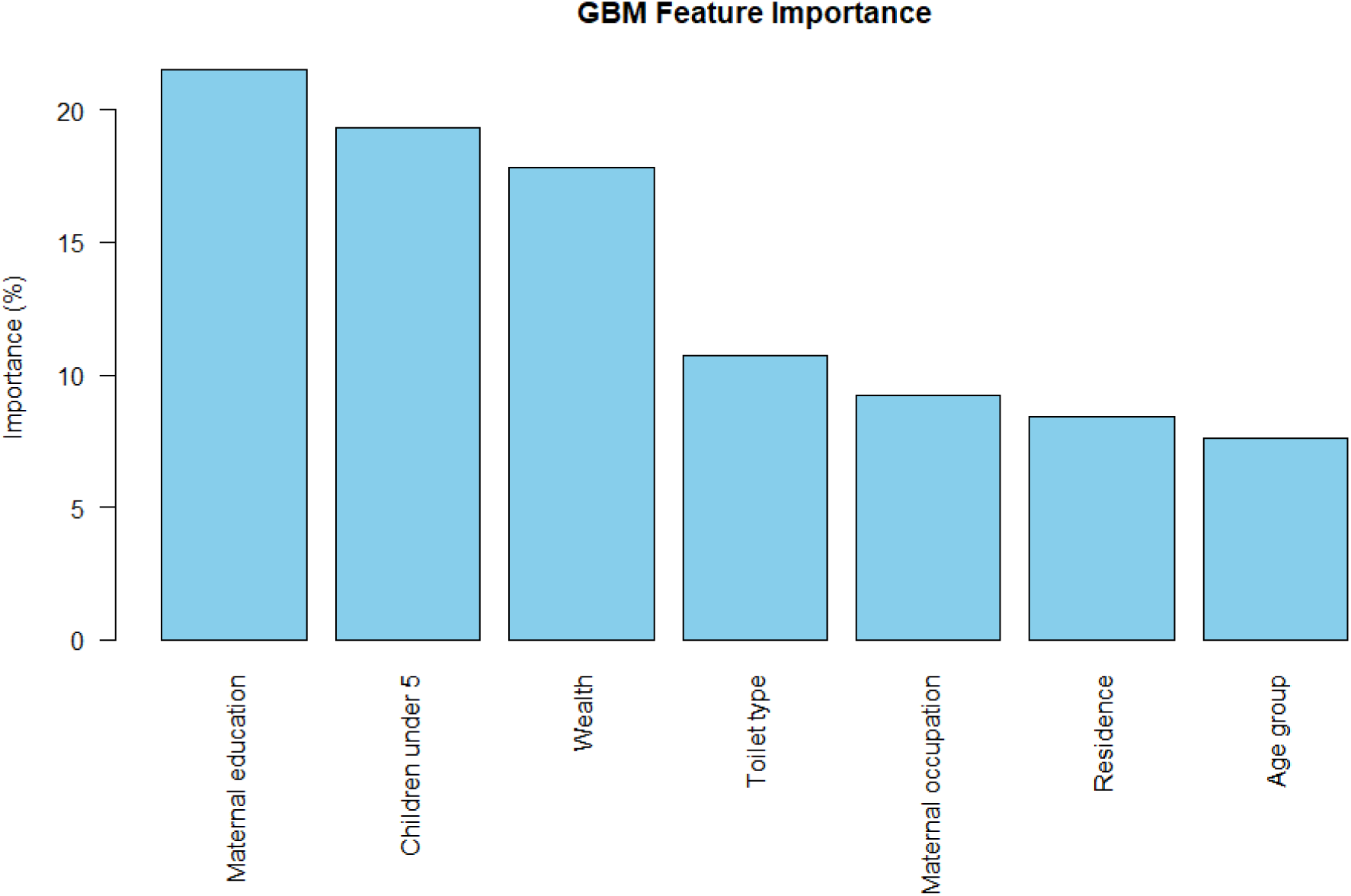
Feature importance for predicting anemia among women, identified using the GBM algorithm.

## Discussion

In this study, we found that anemia affected 23.7% of women, with clear evidence of spatial hotspot in northwestern Lesotho (Leribe) and southwestern districts (Mohale’s Hoek and Quthing), while lower clustering was observed in Qacha’s Nek. The Gradient Boosting Machine demonstrated strong predictive performance (AUC = 87.1%), with good balance between sensitivity (72.5%) and specificity (80.3%). Importantly, maternal education, number of children under five, economic status, sanitation, occupation, residence type, and age emerged as the most influential determinants, underscoring the multifactorial nature of anemia risk in this setting.

In this study, 23.7% of women in Lesotho were found to be anemic, indicating that nearly one in four women is affected. This finding is lower than studies conducted in Ethiopia (56.8%) (20), Nigeria (54.5%) (21), Burkina Faso (54.4%) (22), Uganda (32%) (23) and Ghana (28.7%) (24). In contrast, the prevalence is higher than that reported in The Gambia (21.6%). Variations in anemia prevalence between populations can be influenced by a combination of geographical, socioeconomic, seasonal, nutritional, and behavioral factors (25, 26). For example, differences in climate and regional food availability can affect nutritional intake, while income levels and educational attainment shape access to healthcare and health-related knowledge. Seasonal fluctuations in food supply and disease exposure may also contribute to changing anemia rates throughout the year. Moreover, infections such as malaria and intestinal parasites are important biological determinants, as their presence can directly reduce hemoglobin levels and exacerbate anemia (27).

The spatial distribution of anemia in Lesotho is non-random. A spatial hotspot of anemia was detected in northwestern Lesotho (Leribe) and in the southwestern districts (Mohale’s Hoek and Quthing), while a cold spot was observed in Qacha’s Nek. A study from Congo revealed that the distribution of anemia is random (28), whereas studies from Ethiopia showed a clustered distribution, with hotspots in the eastern and northeastern parts of the country (29). Similarly, a study from Mozambique found a clustered distribution of anemia, with hotspot areas in Nampula, Zambezia, and Sofala (30). In addition, research from Nigeria reported high hotspots of anemia across all northern regions of the country (31). These spatial variations in anemia suggest that targeted interventions are needed in hotspot regions to maximize impact. Non-random and clustered distributions indicate that local factor. Understanding these patterns can help policymakers allocate resources more effectively and design context-specific prevention strategies.

The Gradient Boosting Machine (GBM) demonstrated strong predictive performance for anemia. Its ability to handle complex, non-linear relationships allows it to accurately predict individuals and populations at risk. Other studies have similarly shown that GBM is effective for anemia prediction and classification (32, 33). Accurate prediction enables early identification of high-risk groups. This, in turn, supports timely interventions and more efficient allocation of healthcare resources. Overall, GBM provides a reliable tool for predicting anemia and guiding public health strategies.

Higher maternal education was associated with lower anemia prevalence, supported by studies (34, 35, 36) showing that educated women are more likely to make informed decisions regarding diet, healthcare, and family planning, which reduces the risk of anemia. This suggests that promoting female education could have long-term benefits for maternal health. Women with more children under five were at higher risk, likely due to increased nutritional demands and reduced recovery time between pregnancies, consistent with prior research (37) emphasizing the importance of family planning and maternal health support. Strengthening family planning services could help mitigate this risk. Women from wealthier households had lower anemia prevalence, aligning with evidence (38, 39) that higher socioeconomic status improves access to nutritious food, healthcare services, and overall living conditions, thereby reducing anemia risk. Economic empowerment programs for women could therefore contribute to anemia reduction.

Access to improved sanitation, such as proper toilet facilities, was associated with lower anemia prevalence (40, 41, 42), as better sanitation reduces exposure to infections, including parasitic diseases, that contribute to anemia. Investments in sanitation infrastructure could improve maternal health outcomes. Maternal occupation influenced anemia risk (43, 44), with employment providing economic resources that can improve diet and healthcare access, although certain labor-intensive jobs may increase physical strain and nutritional depletion. Supporting women in the workforce through occupational health programs and nutrition education could reduce anemia risk. Residence also affected anemia prevalence, consistent with evidence that urban women generally have better access to healthcare, nutrition, and sanitation compared to rural women (44), highlighting the need to expand health services and nutrition programs in rural areas. Finally, women aged 35–49 years were more vulnerable to anemia (45, 46), likely due to cumulative nutritional deficiencies, repeated pregnancies, and age-related physiological changes affecting iron absorption and blood production. Age-specific interventions, including iron supplementation, dietary counseling, and routine health monitoring, are essential to reduce anemia risk among older reproductive-age women.

Despite the strengths of the study, some limitations should be noted. The cross-sectional design limits the ability to infer causal relationships between the identified determinants and anemia. Some variables were not available in the dataset, which may have influenced the predictive accuracy of the machine learning models. Reliance on secondary survey data may introduce reporting or measurement biases. Future studies should consider longitudinal designs and incorporate additional nutritional and health-related factors to enhance predictive accuracy and causal understanding.

## Conclusions

Anemia affected nearly one in four women in Lesotho, with clear spatial hotspots in northwestern (Leribe) and southwestern districts (Mohale’s Hoek and Quthing). The Gradient Boosting Machine demonstrated strong predictive performance, showing its usefulness for identifying high-risk populations. Maternal education, number of children under five, economic status, sanitation, maternal occupation, residence type, and age were identified as the most influential determinants, highlighting the multifactorial nature of anemia risk in this population. Women with lower education, poorer households, and larger numbers of young children were particularly vulnerable. Limited access to improved sanitation and healthcare in certain areas further contributed to anemia prevalence. These findings underscore the importance of geographically tailored interventions targeting hotspot areas to maximize impact. In addition, broader programs focused on maternal education, economic empowerment, sanitation, and age-specific nutrition are essential. Implementing such strategies can help reduce anemia prevalence and improve overall maternal health outcomes

## Declaration

### Ethics approval and consent to participate

The 2023/24 Lesotho DHS was conducted in accordance with national and international ethical standards. Ethical approval was obtained from ICF International and the U.S. CDC Institutional Review Board. Informed consent was obtained from all participants, while parental or guardian consent and minor assent were secured for respondents under 18 years. Data were collected anonymously, and no personal identifiers were included in this analysis. All procedures followed strict ethical guidelines to ensure privacy, confidentiality, and minimal risk to participants. Further information is available in the LDHS final report (18).

### Consent for publication

None

### Competing interest

There is no conflict of interest.

### Funding statement

There is no specific funding.

### Data availability

The row data for this study are available in the DHS program (https://dhsprogram.com).

### Authors’ contributions

**TAK:** data analysis, manuscript write-up, editing, originated the idea, formulated the protocol and methodology, managed resources and software. **RNH:** data analysis, manuscript write-up, editing, handled project administration, resources, software, supervision, validation."

## Data Availability

The row data for this study are available in the DHS program (https://dhsprogram.com).

## Acknowledgement

The author is indebted to the DHS program for giving permission to access the dataset.

## Notes

### Competing Interest Statement

The authors have declared no competing interest.

### Author Declarations

The row data for this study are available in the DHS program (https://dhsprogram.com).

